# Time trends and prescribing patterns of opioid drugs in UK primary care patients with non-cancer pain: a retrospective cohort study

**DOI:** 10.1101/2020.04.07.20049015

**Authors:** Meghna Jani, Belay Birlie Yimer, Therese Sheppard, Mark Lunt, William G Dixon

## Abstract

**Background:** The U.S. opioid epidemic has led to similar concerns about prescribed opioids in the U.K. In new users, escalation to more potent and high-dose opioids may contribute to long-term use as well as opioid-related morbidity/mortality. The scale of such escalation is unclear for non-cancer pain. Additionally, physician prescribing behaviour has been described as a key driver of rising opioid prescriptions and long-term opioid use. No studies have investigated the extent to which regions, practices, prescribers, vary in opioid prescribing, whilst accounting for case-mix.

**Methods:** Using a retrospective cohort study we used U.K. primary-care electronic health records from Clinical Practice Research Datalink to: (i)describe prescribing trends between 2006-17 (ii)evaluate the transition of opioid dose and potency in the first 2-years from initial prescription (iii)quantify and identify risk factors for long-term opioid use (iv)quantify the variation of long-term use attributed to region, practice and prescriber, accounting for case-mix and chance variation. Adult patients with a new prescription of an opioid without cancer were included.

**Findings:** 1,968,742 new-users of opioids were identified. Rates of codeine use were highest, increasing five-fold from 2006-2017, reaching up to 2,456 prescriptions/10,000 people/year. Morphine, buprenorphine and oxycodone prescribing rates continued to rise steadily throughout the study period. Of those who started on high (100-200 Morphine Milligram Equivalents [MME]/day) or very high dose opioids (>200 MME/day), 4.9% and 10.3% remained in the same or higher MME/day category throughout 2-years, respectively. Following opioid initiation, 15% became long-term opioid users. In the fully adjusted model, MME at initiation, older-age, social deprivation, fibromyalgia, rheumatological conditions, substance abuse, suicide/self-harm and gabapentinoid use were associated with the highest odds of long-term use. After adjustment for case-mix, the North-West, Yorkshire, South- West; 103 practices (25.6%) and 540 prescribers (3.5%) were associated with a significantly higher risk of long-term use.

**Interpretation:** Patients commenced on high MMEs were more likely to stay in the same state for a subsequent 2-years and were at increased risk of long-term use. In the first UK study evaluating long-term opioid prescribing with adjustment for patient-level characteristics, variation in regions and especially practices and prescribers were observed. Our findings support greater calls for action for reduction in practice and prescriber variation by promoting safe practice in opioid prescribing.

**Funding:** Versus Arthritis and National Institute for Health Research

**Research in Context:** *Evidence before this study:* Drug dependence and deaths due to opioids have led to an opioid-overdose crisis in several countries globally including the US and Canada, and subsequent concerns about overprescribing in the UK. Physician prescribing behaviour has implicated as a key driver of rising opioid prescriptions and long-term opioid use however this needs to be assessed in the context of region, GP practice and individual patients. We searched Pubmed and Google Scholar between January 2005 and November 2019, with the terms “opioid” AND/OR “opiate”, “chronic pain” AND/OR “non-cancer pain”, and UK AND/OR England AND/OR “Great Britain” AND/OR “NHS”. We also reviewed relevant reports from Public Health England and other national bodies. The more recent trends for opioid prescribing have included all prescriptions including those for cancer pain, and those that include primary care UK prescription data for non-cancer indications are several years out of date. No studies evaluated how opioid dose and potency changes over time in individual patients after starting an opioid for the first time to assess escalation or tapering. National variation in opioid prescribing reported thus far has not accounted for patient case-mix. No studies have assessed the effect of the prescriber on opioid prescribing adjusting for regional, practice level variation and for individual characteristics.

*Added value of this study:* There has been a substantial overall increase in opioid-prescribing for non-cancer pain with clear drug-specific trends between 2006-17. To our knowledge, this is the first UK study that has evaluated the sequential transition on how dose/potency vary when a patient is first prescribed an opioid in primary care. Furthermore we report for the first time the effect of individual risk factors, UK regions, GP practice and prescriber (whilst considering these elements together) on long-term opioid use.

*Implications of all the available evidence:* Our study highlights the key subpopulations in a UK primary care setting at risk of developing long-term opioid use and the need for closer monitoring of at risk patients. Marked variation between region, practice and prescribers still exists after adjusting for case-mix warranting evidence-based harmonised opioid prescribing guidelines with clearer MME/day thresholds. On a practice level, guidance on regular review and dose reduction, as well as using prescriber and practice variations as a proxy for quality of care through audit and feedback, to highlight unwarranted variation to prescribers, could help drive safer prescribing.

## INTRODUCTION

The sharp increase in prescription opioid use for non-malignant pain in the U.S., Canada and several European countries^1–3^ has led to concerns of a similar epidemic in the UK. Opioids have now become the leading cause of accidental death and unintentional injury in the U.S.^4^. In the UK, opioid-related deaths have been increasing over the last few decades, the majority of which are non-intentional^3,5,6^. Alongside this, a rise in opioid prescribing using national population level prescribing datasets has been reported (for all indications including cancer)^7,8^. A recent Public Health England analysis revealed 13% of the UK adult population had one or more prescriptions of opioids dispensed between 2017-18^9^.

Opioids are associated with several serious adverse outcomes that are believed to be dose and potency dependent^10^. The escalation rate to higher doses and more potent opioids is likely to also contribute to long-term prescriptions, which in turn may be associated with opioid dependence, addiction and overdose^11^. Until recently, several commonly prescribed opioids did not have a recommended maximum dose, despite minimal evidence of benefit in non-chronic pain at higher doses. This may lead to considerable variation in opioid prescribing in the context of chronic pain following initiation, including transitioning to stronger opioids, higher dose, combination opioids or not reducing dose in a timely manner. The longitudinal opioid pathway of patients commencing opioids for non-cancer pain, the scale of dose escalation/tapering and ensuing long-term use, remains unexplored.

Variation in opioid prescribing across UK regions has been described recently on a population level from Clinical Commissioning Groups^8^. Furthermore, physician prescribing behaviour has been described to be one of the key drivers of rising opioid use^12^. However, the effect of region, practice and prescriber requires interpretation within their context, by accounting for individual patient characteristics. No studies have investigated the extent to which regions, practices and individual general practitioners (GP) vary in opioid prescribing, accounting for the patient (case) mix nor the implications of such variations on long-term opioid prescribing. Identification of what individual patient characteristics are associated with long-term opioid prescribing in primary care would allow prescribers to exercise vigilance and explore alternatives to opioids where appropriate in certain patient subgroups.

The study objectives were to: (i) describe trends of the most commonly prescribed opioids for non-cancer pain in UK primary care over a 12-year period (2006-17) in new users (ii) assess the transition of Morphine Milligram Equivalents [(MME) accounting for dose, opioids type and sequence of use] in the first two-year period after first prescription (iii) quantify and identify risk factors for the transition from new-users to long-term opioid users, and (iv) quantify the variation of long-term use attributed to region, practice and prescriber, accounting for patient mix and chance variation.

## METHODS

### Data source

We conducted a retrospective observational study from 1/1/2006 to 31/12/2017 using the Clinical Practice Research Datalink (CPRD), a database of anonymised UK primary care electronic health records. CPRD is one of the largest research databases of longitudinal primary care records in the world and contains information from >15 million patients. Prescriptions are recorded electronically and clinical information including diagnoses are documented using Read codes.

### Study population

Patients aged ≥18 years without prior cancer who were new users of opioids were identified, in order to establish an incident user cohort prescribed opioid for non-cancer indications. A 24 month ‘wash-out’ period prior to the index date was used to identify new users. Patients with a previous history of a malignancy Read code up to 10-years prior to index date were excluded, with the exception of non-melanoma skin cancer. Follow up start was defined as the date of the first opioid prescription for a given individual (index date). Patients stayed in the cohort until end of follow up, death or if they left the practice. Patients on methadone were excluded because, in the UK, it is primarily prescribed as an opioid addiction treatment and not consistently prescribed by GPs. Supplementary figure 1 describes the derivation of the cohort.

### Covariates

Baseline characteristics such as age, gender, ethnicity, comorbidities that comprise the Charlson comorbidity index^13^, smoking and deprivation scores were measured using data from the year prior to index date. Socio-economic status was assessed using linked-data for Townsend deprivation scores, a measure of material deprivation based on UK Census data^14^.

To identify risk-factors for long-term opioid use, we identified additional *a priori* variables, based on clinical knowledge and published literature. All diagnoses were identified using Read codes 1-year prior to first opioid prescription. The Centre for Disease Control and Prevention (CDC) has identified certain factors associated with opioid misuse such as previous substance-use disorder, major depression and use of psychotropic medications^15^, which we defined in CPRD. Psychotropic medications included antiepileptics, antihistamines, antiparkinsons, antipsychotics, anxiolytics, hypnotics and sedatives. Other factors evaluated included prior history of suicide and self-harm, alcohol excess, major surgery in the last 1-year, and pain conditions such as back pain, migraine and fibromyalgia. Rheumatological disorders included rheumatoid arthritis, systemic lupus erythematosus, myositis and giant cell-arteritis (defined by the Charlson score^13^). The use of psychotropic medications, benzodiazepines and gabapentinoids was defined as any use 1-year prior to the index date, including the date the first opioid was prescribed. Prescriber, GP practice and regional information for each patient were obtained to examine variation at each level in opioid prescribing.

### Opioid drug preparation and exposure

Opioid exposure data were prepared using a drug preparation algorithm published previously^16^. The decisions made to prepare the data are described in Supplementary figure 2. Classes of opioids were divided into weak opioids (codeine, dihydrocodeine, meptazinol), moderate (tramadol, tapentadol) and strong opioids (morphine, oxycodone, fentanyl, buprenorphine, diamorphine, hydromorphone, pethidine). Tramadol and tapentadol were classed as moderate strength opioids, as despite their low MME they are phenotypically distinct from conventional weak opioids due to their dual mechanism as a partial serotonin-norepinephrine reuptake inhibitor. Combination formulations such as co-codamol, were classed according to their active opioid ingredient. If patients were on ≥1 opioid at index, we categorised them into a separate combination drug group.

To allow direct comparison of doses and opioid potencies across different drugs and formulations we calculated MME for each prescription. MME/day was defined as the daily dose for each prescription multiplied by the equivalent analgesic ratio as specified by the CDC^15^. For transdermal buprenorphine and fentanyl formulations, strength per hour and the duration of delivery rate of the formulation was considered in the dose calculation to avoid underestimation of daily MME. An episode of long-term opioid use was defined as at least three opioid prescriptions issued within a 90-day period from index date, or ≥1 opioid prescription lasting at least 90 days in the first year of follow-up. When defining long-term use, we ignored the first 30-day period following index date to allow for acute pain treatment (including any 90-day period after day 30 as defined above up to 1-year as long-term use).

### Statistical analysis

Descriptive statistics were used to assess the baseline characteristics of the cohort, stratified according to opioid strength at initiation.

#### Prescribing trends over time

To evaluate trends of opioid prescribing over time, the rate of prescriptions for each opioid drug was calculated by calendar year by dividing the number of prescriptions per year for the cohort (numerator) by the number of eligible patients registered in CPRD per year (denominator). Raw denominator figures of patients registered were provided by CPRD in April 2018 and prepared for use (Supplementary Material).

#### Transition of opioids over two years

Patients were stratified into four categories according to the average MME/day in the first three months after index date to incorporate the type, potency and dose of the opioid. MME categories were as follows: low, <50MME/day; medium 50- 100MME/day; high, 100-200MME/day and very high >200MME/day. For instance a prescription of 30mg codeine four times a day would equal 18 MME/day. An oxycodone prescription of 40mg four times/day equates to 240 MME/day. In patients on combination of opioids, MME was calculated for each drug and the sum was taken as the MME/day. Sunburst plots, as described previously^17^, were created to quantify visually the sequential transition of MME/day, in three-month bands, over a two year time window from index date. One plot was generated for each of the MME dosage categories derived from the first three months’ exposure (low, medium, high and very high).

#### Transition from new-users to long-term opioid users

A multi-level random-effects logistic regression model was used to examine the association of different patient characteristics with the odds of becoming a long-term opioid user. Person-level characteristics investigated include age, gender, ethnicity, deprivation scores and comorbidities outlined above. To examine opioid variation amongst prescribers, GP practices and region after adjusting for patient case-mix, we used a nested random effect structure (i.e., prescribers nested within practices and practices nested within region). The approach introduced by Snijders and Bosker^18^ was followed to obtain the explained variation at each level of the hierarchy. Furthermore, the posterior distributions of the prescriber, practice and region level random effects were simulated using the REsim function in the merTools package^19^ for the fully adjusted models. The adjusted random-effect estimates along with 95% confidence intervals were then ranked and plotted on an odds ratio (OR) scale as well as percentage value. Those with the lower end of the 95% confidence intervals (CI) >1 were associated with a higher risk, and those with the upper end <1 with a lower risk of long-term opioid use. That is, a ‘high-risk’ prescriber or practice was defined as those prescribers or practices where the entire adjusted 95% CI lay above the population average (i.e. 1). A ‘high-risk’ region, practice or prescriber were defined as those where the entire adjusted 95% CI lay above the population average (i.e. 1). The risk of becoming a long-term opioid user attributed to a specific practice was then plotted against the proportion of high-risk prescribers within each practice to evaluate the influence of a high-risk prescriber on the practice (supplementary material). The study was approved by the CPRD’s Independent Scientific Advisory Committee (approval number: 16_278). All analyses were performed in STATA version 14.0 and R version 3.5.0.

### Role of the funding source

The funders had no role in study design, data collection, data analysis, data interpretation, writing of the report, or the decision to submit for publication. All authors had full access to all the data in the study and had final responsibility for the decision to submit for publication.

## RESULTS

We identified 1,968,742 opioid new users who met our inclusion criteria, of which 88.2% were initially commenced on a weak opioid, 8.5% on a moderate, 2.6% on a strong opioid and 0.7% on combination drugs (Table 1). The highest proportion of new opioid users for the weak, moderate and combination categories were between 35-54 years, whereas patients who were started on strong opioids were older: 31.5% of strong opioids were prescribed to patients ≥85 years (compared to 4.1% and 3.3% in the weak and moderate opioid groups respectively). Proportionally more patients commencing strong opioids had higher Charlson scores of≥4 (7.5% compared to <2% in other opioid groups). Townsend deprivation quintiles were represented in similar proportions across weak, medium, strong and combination opioids. The proportion of patients on all types of opioids were slightly lower in the most deprived category, between 11-16%. The strong opioid group had the highest proportions of patients on prior benzodiazepine, gabapentinoid or psychotropic medications (Table 1).

**Table 1:** Baseline characteristics of opioid new users grouped by opioid at initiation.

| Characteristics<br>N = 1,968,742 | Monotherapy at index |  |  | Combination therapy<br>at index |
| --- | --- | --- | --- | --- |
|  | Weak opioids<br>(n = 1,736,398)<br>88.2% | Moderate opioids<br>(n = 166,524)<br>8.5% | Strong opioids<br>(n = 51,126)<br>2.6% | Combination opioids<br>(n = 14,694)<br>0.7% |
| Age, years (mean, SD) | 50.5 (19.0) | 51.1 (17.6) | 69.6 (21.0) | 54.9 (19.9%) |
| **Age group (years), n (%)** |  |  |  |  |
| 18-24 | 155,350 (9.0) | 10,175 (6.1) | 1,061 (2.1) | 739 (5.0) |
| 25-34 | 262,288 (15.1) | 22,238 (13.4) | 3,060 (6.0) | 1,778 (12.1) |
| 35-44 | 300,793 (17.3) | 31,705 (19.0) | 4,089 (8.0) | 2,525 (17.2) |
| 45-54 | 300,381 (17.3) | 33,763 (20.3) | 4,942 (9.7) | 2,715 (18.5) |
| 55-64 | 273,806 (15.8) | 28,946 (17.4) | 5,418 (10.6) | 2,314 (15.8) |
| 65-74 | 218,957 (12.6) | 20,662 (12.4) | 6,228 (12.2) | 1,773 (12.1) |
| 75-84 | 153,738 (8.9) | 13,515 (8.1) | 10,206 (20.0) | 1,341 (9.1) |
| ≥85 | 71,085 (4.1) | 5,520 (3.3) | 16,122 (31.5) | 1,509 (10.3) |
| Female, n (%) | 991,322 (57.1) | 92,675 (55.7) | 31,913 (62.4) | 8,178 (55.7) |
| **Ethnicity, where reported\*, n (%)** |  |  |  |  |
| White | 1,114,790 (89.0) | 115,023 (91.4) | 37,116 (94.6) | 10,016 (92.1) |
| Asian | 51,499 (4.1) | 3,479 (2.8) | 596 (1.5) | 259 (2.4) |
| Black | 29,231 (2.3) | 1,983 (1.6) | 307 (0.8) | 144 (1.3) |
| Other | 18,979 (1.5) | 1,437 (1.1) | 266 (0.7) | 111 (1.0) |
| Mixed | 10,794 (0.9) | 892 (0.7) | 164 (0.4) | 71 (0.7) |
| Unknown | 27,812 (2.2) | 2,998 (2.4) | 765 (2.0) | 276 (2.5) |
| **Townsend Deprivation score, where reported \*, n (%)** |  |  |  |  |
| Q1 (least deprived) | 193,601 (20.8) | 19,180 (21.0) | 6,539 (21.9) | 1,632 (21.6) |
| Q2 | 194,478 (20.9) | 20,320 (22.2) | 7,651 (25.6) | 1,705 (22.5) |
| Q3 | 194,884 (21.0) | 19,004 (20.8) | 6,729 (22.5) | 1,630 (21.5) |
| Q4 | 198,714 (21.4) | 19,285 (21.1) | 5,766 (19.3) | 1,565 (20.7) |
| Q5 | 147,054 (15.8) | 13,599 (14.9) | 3,182 (10.7) | 1,041 (13.7) |
| **Charlson comorbidity score, n (%)** |  |  |  |  |
| • Low score (0) | 1,249,477 (72.0) | 117,614 (70.6) | 25,101 (49.1) | 9,732 (66.2) |
| • Medium score (1-3) | 454,906 (26.2) | 45,652 (27.4) | 22,181 (43.4) | 4,527 (30.8) |
| • High score (≥4) | 32,015 (1.8) | 3,258 (2.0) | 3,844 (7.5) | 435 (3.0) |
| **Conditions associated with opioid dependency** |  |  |  |  |
| History of alcohol dependency, n (%) | 37,787 (2.2) | 4,496 (2.7) | 1,162 (2.3) | 452 (3.1) |
| History of substance use disorder, n (%) | 25,851 (1.5) | 3,558 (2.1) | 1,702 (3.3) | 374 (2.6) |
| History of depression, n (%) | 374,209 (21.6) | 40,350 (24.2) | 10,756 (21.0) | 3,440 (23.4) |
| History of suicide & self-harm, n (%) | 57,119 (3.3) | 7,265 (4.4) | 1,834 (3.6) | 815 (5.6) |
| **Concomitant medication, n (%)** |  |  |  |  |
| Baseline benzodiazepine use † | 156,050 (9.0) | 20,252 (12.2) | 14,319 (28.0) | 2,806 (19.1) |
| Baseline gabapentinoid use † | 22,627 (1.3) | 6,495 (3.9) | 4,054 (7.9) | 1,235 (8.4) |
| Baseline psychotropic drug use † | 197,133 (11.4) | 24,996 (15.0) | 18,752 (36.7) | 3,654 (24.9) |
| **Other** |  |  |  |  |
| Major surgery n (%) | 41,088 (2.4) | 10,490 (6.3) | 2,354 (4.6) | 1,231 (8.4) |
| **Smoking status, where reported\*, n (%)** |  |  |  |  |
| Never | 341,075 (45.5) | 30,060 (42.3) | 9,708 (49.2) | 2,601 (44.0) |
| Former | 220,141 (29.4) | 21,392 (30.1) | 6,614 (33.5) | 1,672 (28.3) |
| Current | 188,185 (25.1) | 19,671 (27.7) | 3,423 (17.3) | 1,639 (27.7) |
| **\*Missing, n (%)** |  |  |  |  |
| Ethnicity | 483,293 (27.8) | 40,712 (24.5) | 11,912 (23.3) | 3,817 (26.0) |
| Townsend Deprivation score | 807,667 (46.5) | 75,136 (45.1) | 21,259 (41.6) | 7,121 (48.5) |
| Smoking status | 986,997 (56.8) | 95,401 (57.3) | 31,381 (61.4) | 8,782 (59.8) |

NB: Proportions are presented as percentage of non-missing data. †Baseline use refers to use 1-year prior and including index date. Abbreviations: Q1, Quintile; SD, standard deviation

### Population level prescribing opioid trends

The most commonly used opioids were codeine, dihydrocodeine and tramadol. Over a 12-year period between 2006-2017, codeine use increased fivefold from 484 to 2456 prescriptions per 10,000 population/year. Dihydrocodeine, tramadol and fentanyl prescriptions increased between 2006-2012, and plateaued thereafter until end of 2017. Within the strong opioids group, oxycodone prescribing rose ∼30-fold from 5 to 169 prescriptions per 10,000 population over 12 years. Morphine prescriptions also rose considerably from 18 to 422 prescriptions per 10,000 population/year between 2006 and 2017 (Figure 1).

**Figure 1:**
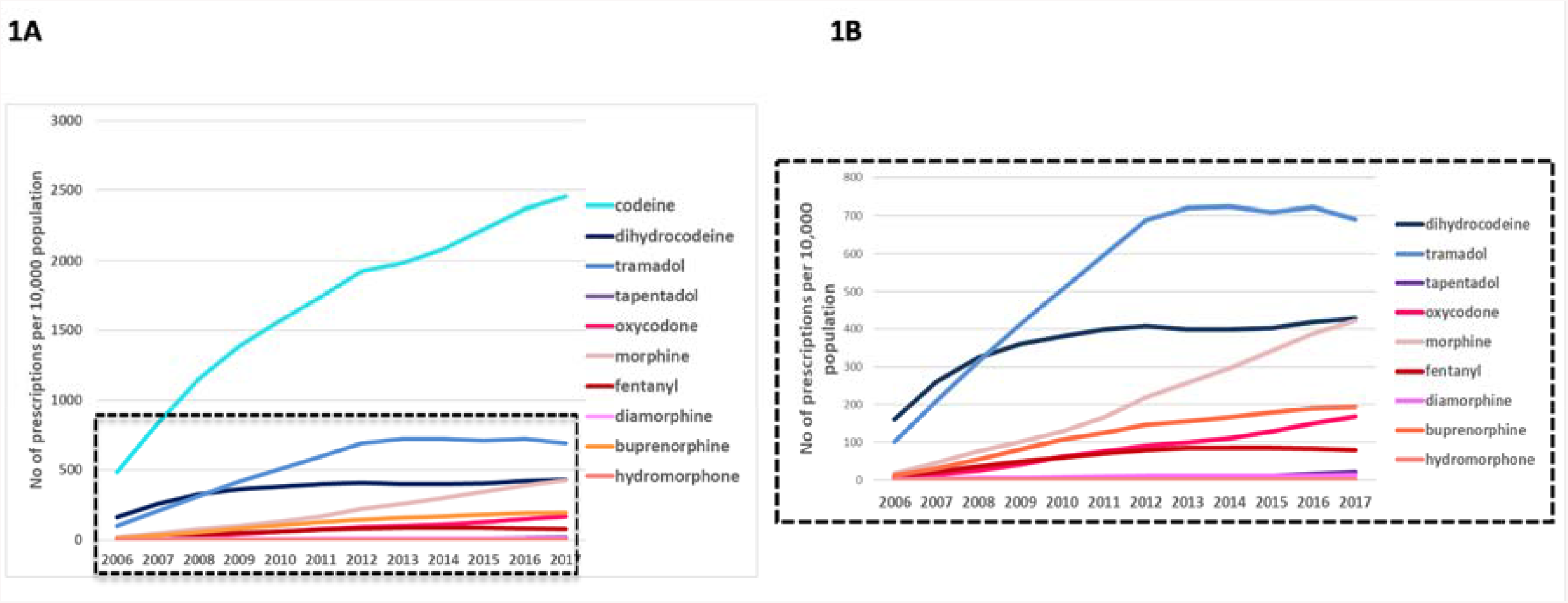
Trends in opioid utilisation in CPRD by individual opioi. Figure 1A represents the most frequently prescribed opioids between 2006-2017. Figure 1B represents all opioids except for codeine. Weak/moderate strength opioids are represented in shades of blue and strong opioids in shades of red.

The transition of MME state in new users of opioids over 2-year period is shown in Figure 2. The majority of patients were started on MME <50/day (n=1,925,944, 98.5%), of whom 1,210,973 (62.9%) stopped completely over the 2 -years and 714,971 (37.1%) continued in the same state or higher over the 2 years. Less than 1.5% of patients who started on a low MME (<50), transitioned to higher MME categories over 2 years. At 6 months 6,007 (0.3%), at 1 year 13,580 (0.7%) and at 2 years, 24,231 (1.3%) transitioned to a medium (50-100), high (100-200) or very high (>200) state, respectively. Of the 22,397 (1.2%) patients who started on a medium MME between 50-100/day as their starting dose, by 1 year 1,133 (5.1%) escalated to the higher MME group, whilst 16,613 (74.2%) were tapered to <50 MME/day (including discontinuation). By 2-years 1,315 (5.9%) escalated to the high MME group, 306 (1.3 %) to the very high MME group and 16,165 (72.2%) were tapered to the low MME group or discontinued. Patients commencing on very high MME of >200/day (n=1,446 [0.08%]) were more likely to stay on in a higher MME category throughout the 2 years. Of the 1,446 patients, 656 (45.4 %) continued to be on very high MME/day throughout the 6 months, 334 (23.1%) throughout 1 year and 148 (10.3%) throughout 2 years. By 2 years 96 patients (6.6%) tapered down to an MME of 100-200/day, 28 (1.9%) to 50-100 MME/day and 32 (2.2%) to <50MME/day.

**Figure 2:**
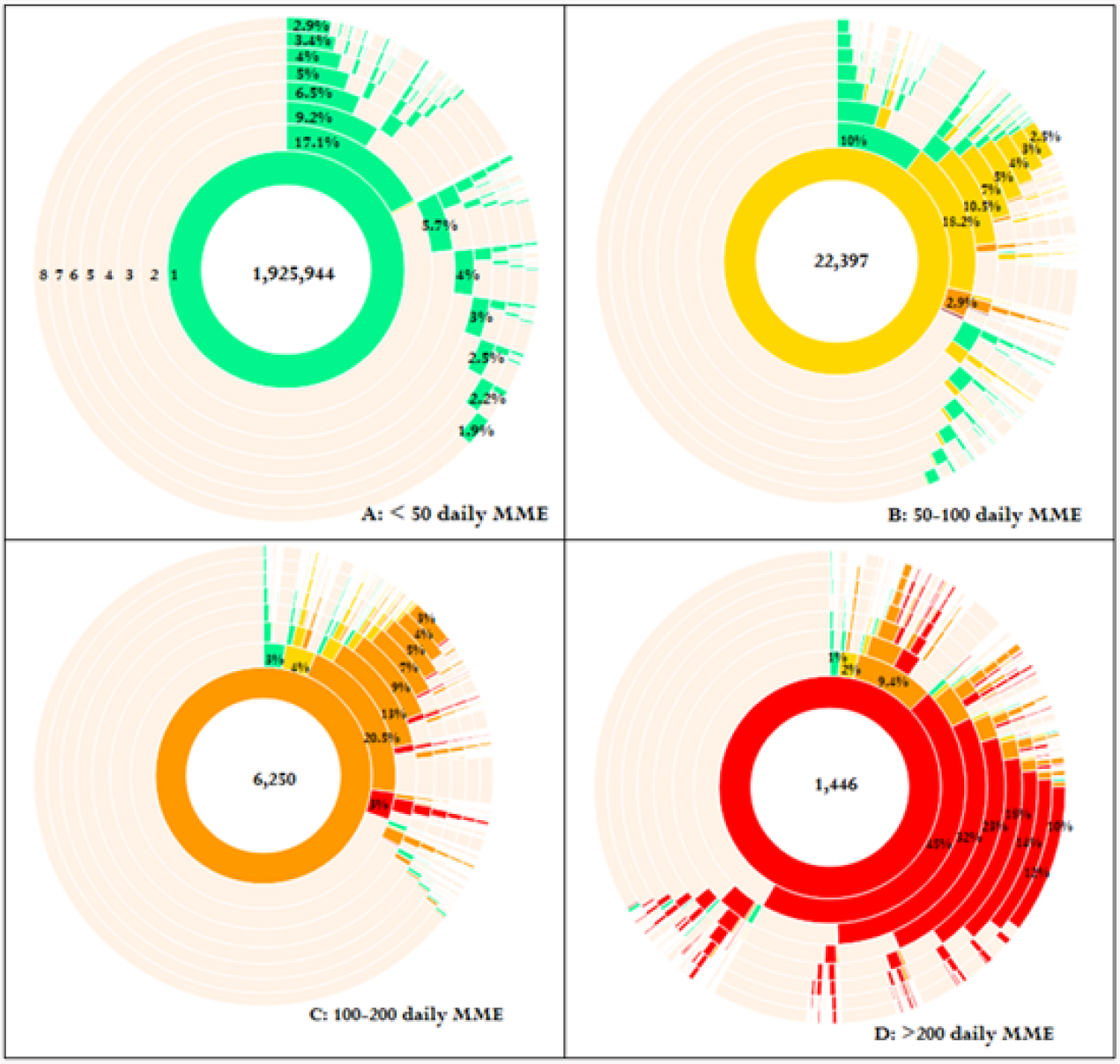
Sunburst diagram demonstrating progression of MME dosage categories over 2-year period from index date, stratified by dose categories in the first three months. The inner circle demonstrates the average MME state patients are exposed to during the first 3 months, the second circle shows the average MME level the patients are exposed to during the second 3 months, and so forth. A: Patients started on <50 MME/day; B: 50-100 MME/day; C: 100-200 MME/day D: >200 MME/day. Green indicates <50 MME/day; yellow, 50-100 MME/day; orange, 100- 200 MME/day; red, >200MME/day.

### Variation of long-term opioid use by prescriber, practice and region

In our new user cohort, 14.6% became long-term opioid users in the first-year post-index date. In the fully adjusted model, a number of individual factors were identified to be associated with a higher odds of long-term opioid use including older age, social deprivation, fibromyalgia, suicide/self-harm, excess alcohol, previous gabapentinoid use, psychotropic use and major surgery and initial dose(Figure 3). The strongest association was seen in those who were prescribed >200 daily MME at index, who were 7.6 (95% CI 6.3-9.2) times more likely to become a long-term opioid user compared to those who started on <50 daily MME/day.

**Figure 3:**
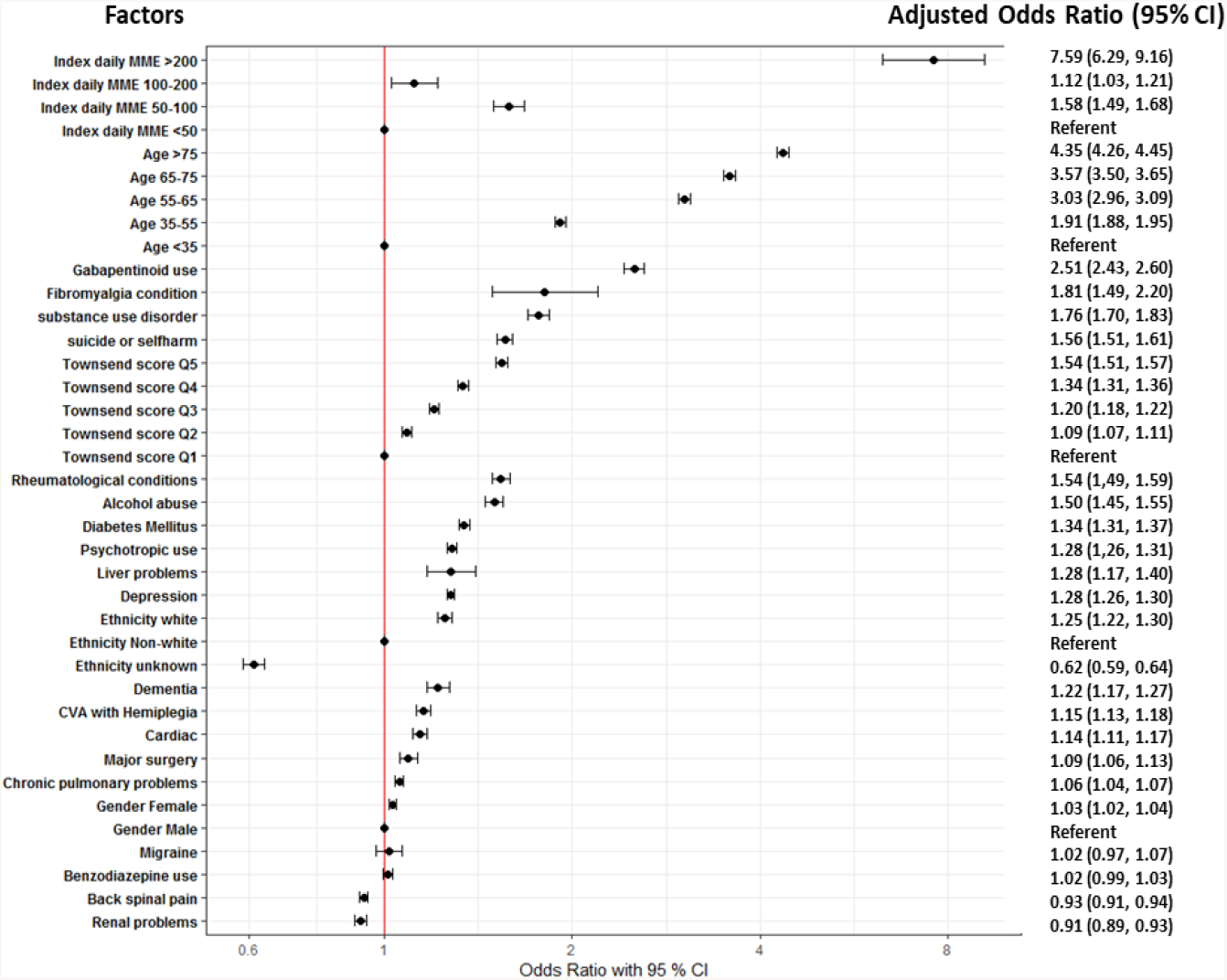
Factors associated with long term opioid use using a multi-level model accounting for clustering of individuals within prescriber, practice and region.

Figure 4 illustrates the prescriber, practice and regional level variation in the odds of long-term opioid use. After adjustment for case-mix, there was still considerable variation among regions, practices and prescribers. Prescribers explained 2.2% of the total variation whilst practices and regions explained 0.6% and 0.01% of the variation, respectively. Three regions (namely North-West, Yorkshire & the Humber, and South-West) had proportions of long-term opioid users signficantly greater than the population average (Figure 4A). The proportion of long-term users for North- west, Yorkshire & the Humber, and South-west was 16% (95% CI: 15, 17), 15% (95% CI: 15, 16), and 15% (95% CI: 15, 16), respectively while the proportion of long-term users in London was 13% (95% CI: 12, 13). High levels of variation were observed among practices after case-mix adjustment. Of the 402 practices included in the study, 103 practices (25.6%) were associated with a significantly higher risk of long-term opioid use(Figure 4B). The proportion of long-term users for the most high-risk practice was 23% (95% CI: 19, 28) (OR 1.79, 95% CI: 1.42-2.26), while the proportion of long-term users for the least at risk practice was 10% (95% CI: 9, 11) (OR 0.60, 95% CI: 0.53-0.68). (Supplementary material Figure 3). After case-mix adjustment, 540 (3.5%) prescribers were associated with a significantly higher risk of long-term opioid use. This small proportion of high-risk prescribers had notably high prescribing behaviour, with the odds of becoming a long term user reaching up to an OR of 3.56 (2.53, 5.02) times compared to the population average (Figure 4C). This equated to around 37% of new users for the highest risk prescriber became long-term users by the end of the first year. In certain high-risk practices, the propensity of becoming high-risk was driven by a few prescribers (Supplementary material Figure 4).

**Figure 4:**
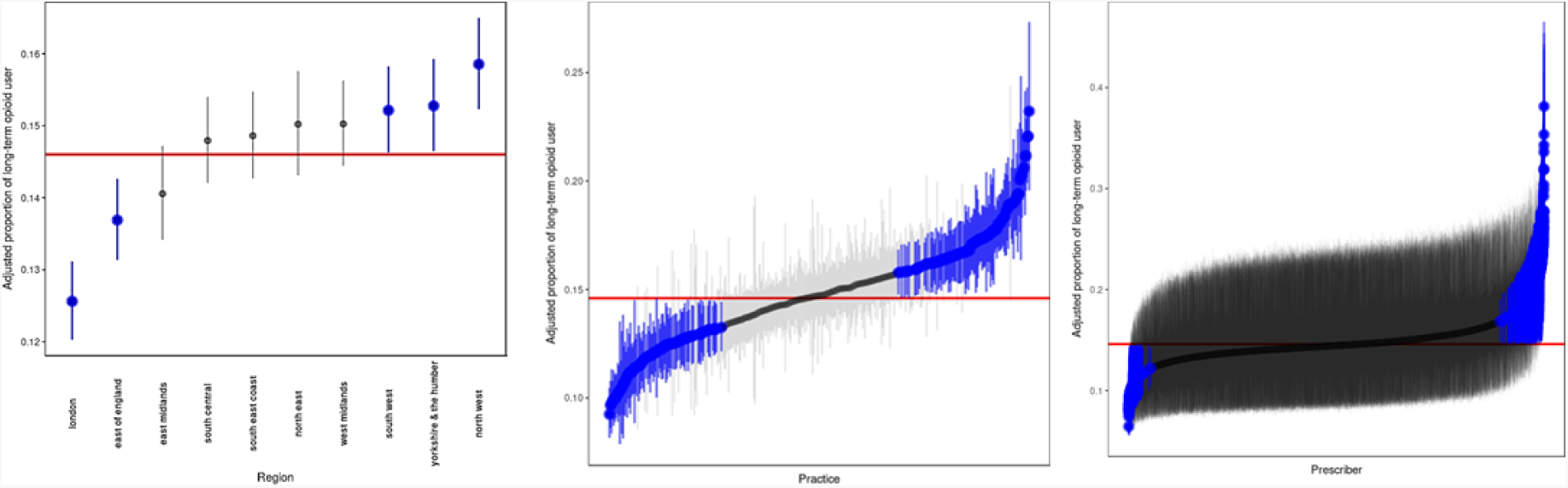
Level of variation among regions (A), practices (B) and prescribers (C) in terms of the odds of long term opioid use. Each vertical line represents the point estimate with 95% confidence intervals with region or practice or prescribers. Regions, practices or prescribers with 95% confidence intervals entirely above or below the population average are indicated in blue. For instance, the adjusted proportion of long-term users for North-west region (15.8%) is significantly higher than the population average (14.6%). The largest variation is seen among practices and prescribers. The proportion of long-term opioid users for some practices reaches up to 23.2%. The proportion of long-term opioid users for the highest risk prescriber is 37.2%.

## DISCUSSION

In this large national cohort of opioid naïve patients in CPRD, we found a substantial increase in opioid prescribing for non-cancer pain between 2006-2017. Of the patients who started on high (100-200 MME/day) or very high dose opioids (>200 MME/day), 15.1% and 30.0% remained in the same or higher MME/day category throughout 1-year respectively. We identified a number of patient-specific factors associated with long-term opioid not previously identified in a UK population, most notably high initial dose/potency of opioid, fibromyalgia, rheumatological conditions, history of depression, prior gabapentinoid/psychotropic use and history of major surgery. A wide variation in the risk of long-term opioid use was observed by prescriber, practice and region. In addition, a regional divide in long-term opioid use risk was found, with the North-West, Yorkshire and South-West associated with the highest level of long-term opioid use. Whilst there was a small proportion of prescribers (3.5%) that had significantly higher prescribing practices, where they did, they were considerably higher in comparison to the population average. After adjusting for case-mix, certain prescribers within a practice could be observed to be driving their entire practice towards high long-term opioid prescribing.

### Comparison with previous studies and interpretation

To our knowledge this is the largest UK study evaluating opioid prescribing trends for non-cancer pain, with patient-level data to ascertain total amount of drug prescribed in terms of MME/day. We addressed a number of key questions quantifying the variance in prescribing at the regional, practice and prescriber level. The overall rise in opioid prescribing trends are complementary to a recent study using NHS digital pharmacy claims data demonstrating 34% increase in opioid prescriptions between 1998-2016^7^. This study however included prescription data on all opioids including cancer, lacked individual-level data and high dose MME definitions were based on presumptions of daily dose. Our results are consistent with a previous cross-sectional CPRD study that reported a rise in morphine, oxycodone, fentanyl and buprenorphine prescribing between 2000-2010 in a non-cancer population that we were able to extend both in time-frame and other opioids^20^. In our study, codeine, morphine and buprenorphine prescriptions in particular continued to rise until end of 2017. Since 2013 in the UK, national regulations have been designed to improve the use and monitoring of controlled drugs such as opioids^21^. An increase in tramadol, oxycodone and fentanyl prescriptions continued until 2012 following which prescribing plateaued, suggesting that GPs may have already started to reduce use of new opioids for these medications earlier. However, such regulations did not change prescribing trends for morphine or buprenorphine.

Clinicians have an opportunity to be vigilant about what type of patient may become a long-term opioid user. A number of individual features associated with increased odds of long-term opioid use were identified. Older age and social deprivation were associated with an incremental increase in risk of long-term opioid use (Figure 3). Clinical Commissioning Group level deprivation has been associated with higher population-level opioid prescribing using NHS digital data^8^. In the U.S, substance abuse, depression and psychotropic medicines have been associated with an increased risk of opioid-misuse^15,22^, and we found these factors to also be associated with an increased risk of long-term opioid use in opioid naïve patients in the UK. Additionally, benzodiazepines and gabapentinoid use, also significantly increased odds and may be a surrogate for chronic pain severity. Concomitant use with opioids have also been associated with an increased risk of death^23,24^. Additionally a history of specific conditions such as alcohol excess, fibromyalgia, rheumatological conditions, diabetes and prior major surgery were significantly associated with a higher odds of long-term opioid use. New opioid users especially post-surgery in the U.S., have been shown to be a vulnerable population both for new persistent use and for developing opioid dependence/overdose^25,26^. Therefore, addressable patient-level factors and certain vulnerable groups at a higher risk of long-term use warrants increased awareness in prescribing clinicians.

An important finding in our study was that 14.6% of new users became long-term users over a 1-year period. In patients who were started on high doses of opioids (>100 MME/day), a considerable proportion continued on higher doses throughout the following year. We also observed a wide variation at a practice and prescriber level in the adjusted odds of becoming a long-term opioid user (Figure 4 B/C) and the propensity of a practice being ‘high risk’ for its patients becoming long-term opioid users was being driven by a few prescribers in some cases. There are a few possibilities why prolonged opioid use may occur, in addition to ongoing appropriate prescribing for patients with clinical need. The variability in prescribing may in part be explained by unclear guidance regarding best practice in managing non-cancer pain. The advice regarding MME/day thresholds beyond which tapering should occur varies internationally^15,27^, therefore GPs may not be aware of which patients to intervene with. In the U.S. national guidelines advise precautions and reassessment of patients exceeding 50 MME per day, and should avoid increasing dose to 90 MME/day or more per day ^15^. The Faculty of Pain Medicine in the UK, suggests harms outweigh benefits when patients exceed over 120 MME/day^27^. There is minimal guidance based on scientific evidence on how best to reduce/discontinue opioids in chronic pain. Clinicians may be guided by patients, who may understandably fear worsening pain, withdrawal symptoms, lack adequate social/healthcare support or could perceive a lack of effectiveness of non-opioid pain relief options^28^. Alternatively transition to long-term opioid use could be driven by “clinical inertia” in some instances^29^, where prescribers continue providing repeat prescriptions, assuming drug effectiveness without regular review.

The adjusted odds for long-term opioid use in opioid naïve patients was highest in the North-West, Yorkshire and South-West of England (Figure 4A). A regional UK variation in population level opioid prescribing between the North and the South, has been observed in recent studies^7,8^. A previous study using NHS digital data showed that nine out of 10 of the highest prescribing areas in the country, were located in the North of England and there was an association with social deprivation^8^. Health is known to be worse in the North and a strength of this study was that we were able to account for case-mix also, while previous studies have not. Whilst chronic pain severity was not measured, there is no known significant regional variation in the prevalence of chronic pain across strategic health authorities^30^. Therefore, it should not account for the observed regional differences in long-term opioid prescribing. We found even after adjusting for deprivation indices (which has been linked to chronic pain^30^), the North/South disparities in long-term opioid users continued to exist.

### Limitations of this study

In the UK, codeine and dihydrocodeine were available over the counter during the study period. Because CPRD data captures electronic prescription data from primary care physicians, the findings likely underrepresent overall drug utilisation of weaker opioids. In 2014 tramadol was reclassified as a schedule 3 drug and prescriptions longer than one month prohibited at any one time. Therefore, the rise in prescriptions may reflect shorter prescriptions for certain medications. The advantage of using this measure is it allows for comparisons to other studies internationally.

Opioid exposure is complicated by the possibility that patients may not fill their prescriptions, administered by the patient, can be taken as required or there may be issues around divergence, none of which are captured in primary care databases. Since data for this study were collected as part of routine-clinical care, we did not have access to patient level pain scores, disease severity of underlying diseases or patient perceptions of opioid prescribing. We did however adjust for benzodiazepine, gabapentinoid and psychotropic drug use that could be used as a potential proxy for pain severity.

## Conclusions

In conclusion, overall opioid prescribing has increased over 12 years with wide variation across the UK after adjusting for patient characteristics. There were considerable differences within practices/prescribers in opioid prescribing and the associated risk of long-term opioid use, even after adjusting for case mix. Patients started on high MMEs were more likely to stay in the same state for the following two years. Whilst reasons are likely to be multi-factorial, exercising vigilance when prescribing to those with identified individual risk factors and improved educational interventions to improve clinical decision making is likely to be beneficial. Our findings are important as they offer a potential lever for prescribing behaviour change and intervention in subgroups of patients at higher risk of long-term use. On a practice level, guidance on regular review and dose reduction, as well as using prescriber and practice variations as a proxy for quality of care through audit and feedback, to highlight unwarranted variation to prescribers, could help drive safer prescribing.

## Data Availability

No additional data available

## CONTRIBUTORS

MJ and WGD conceived the ideas for the study. MJ, BBY and TS performed the analysis. MJ wrote the first draft of the manuscript. ML provided statistical expertise. MJ, TS, BBY, ML and WGD critically reviewed and edited the manuscript. MJ, BBY, TS, ML and WGD are guarantors of the data and analysis. MJ attests that all listed authors meet authorship criteria and no others meeting the criteria have been omitted.

## DECLARATION OF INTERESTS

All authors have completed the ICMJE uniform disclosure form at www.icmje.org/coi_disclosure.pdf. MJ is a member of the Medicines and Healthcare products Regulatory Agency (MHRA) Opioids Expert Working Group. WGD has received consultancy fees from Google and Bayer, unrelated to this work. All other authors declare no support from any organization for the submitted work; no financial relationships with any organizations that might have an interest in the submitted work in the previous three years; no other relationships or activities that could appear to have influenced the submitted work.

## DATA SHARING

No additional data available.

## ACKNOWLEDGEMENTS

No specific funding was received for directly for this work. This work was supported by the Centre for Epidemiology Versus Arthritis: grant number 20380. M.J.’s work was supported by an NIHR academic clinical lectureship and a Presidential Fellowship. The views expressed are those of the authors and not necessarily those of the NHS, the NIHR or the Department of Health.

## REFERENCES

1 Smolina K, Gladstone E, Morgan SG. Determinants of trends in prescription opioid use in British Columbia, Canada, 2005-2013. Pharmacoepidemiol Drug Saf 2016; 25: 553–9.

2 Boudreau D, Von Korff M, Rutter CM, et al. Trends in long-term opioid therapy for chronic non-cancer pain. Pharmacoepidemiol Drug Saf 2009; 18: 1166–75.

3 Jani M, Dixon WG. Opioids are not just an American problem. BMJ 2017; 5514: j5514.

4 Rudd R, Aleshire A, Zibbell J, Gladden M. Increases in Drug and Opioid Overdose Deaths — United States, 2000–2014. Morb Mortal Wkly Rep 2016; 64: 1378–82.

5 Office for National Statistics. Deaths related to drug poisoning in England and Wales - Office for National Statistics. Stat. Bull. 2017. https://www.ons.gov.uk/peoplepopulationandcommunity/birthsdeathsandmarriages/deaths/bulletins/deathsrelatedtodrugpoisoninginenglandandwales/2016registrations (accessed April 23, 2018).

6 Office for National Statistics. Opioid drug deaths by cause, 1993 to 2015 - Office for National Statistics. 2015. https://www.ons.gov.uk/peoplepopulationandcommunity/healthandsocialcare/drugusealcoholandsmoking/adhocs/0061BQZKqdp2CV3QV5nUEsqSg1ygegLmqRygj015 (accessed July 22, 2019).

7 Curtis HJ, Croker R, Walker AJ, Richards GC, Quinlan J, Goldacre B. Opioid prescribing trends and geographical variation in England, 1998–2018: a retrospective database study. The Lancet Psychiatry 2019; 6: 140–50.

8 Mordecai L, Reynolds C, Donaldson LJ, Williams AC de C. Patterns of regional variation of opioid prescribing in primary care in England: a retrospective observational study. Br J Gen Pr 2018; : bjgp18X695057.

9 Taylor S, Annand F, Burkinshaw P, et al. Dependence and withdrawal associated with some prescribed medicines. Public Health England. London. 2019.

10 Gomes T, Mamdani MM, Dhalla IA, Paterson JM, Juurlink DN. Opioid Dose and Drug-Related Mortality in Patients With Nonmalignant Pain. Arch Intern Med 2011; 171: 686–91.

11 Dunn KM, Saunders KW, Rutter CM, et al. Opioid Prescriptions for Chronic Pain and Overdose. Ann Intern Med 2010; 152: 85.

12 NHS accused of fuelling rise in opioid addiction - BBC News. http://www.bbc.co.uk/news/uk-england-43304375 (xaccessed April 23, 2018).

13 Charlson ME, Pompei P, Ales KL, MacKenzie CR. A new method of classifying prognostic comorbidity in longitudinal studies: development and validation. J Chronic Dis 1987; 40: 373–83.

14 UK Data Service CD. 2011 UK Townsend Deprivation Scores | UK Data Service | Census Data. 2011. https://www.statistics.digitalresources.jisc.ac.uk/dataset/2011-uk-townsend-deprivation-scores (xaccessed July 19, 2019).

15 Dowell D, Haegerich TM, Chou R. CDC Guideline for Prescribing Opioids for Chronic Pain—United States, 2016. JAMA 2016; published online March 15. DOI:10.1001/jama.2016.1464.

16 Pye SR, Sheppard T, Joseph RM, et al. Assumptions made when preparing drug exposure data for analysis have an impact on results: An unreported step in pharmacoepidemiology studies. Pharmacoepidemiol. Drug Saf. 2018; : 781–8.

17 Hripcsak G, Ryan PB, Duke JD, et al. Characterizing treatment pathways at scale using the OHDSI network. Proc Natl Acad Sci 2016; 113: 7329–36.

18 Snijders TAB, Bosker RJ. Multilevel Analysis: An Introduction to Basic and Advanced Multilevel Modeling, 2nd Editio. 2012.

19 Knowles J, Frederick C. Tools for Analyzing Mixed Effect Regression Models [R package merTools version 0.5.0]. 2016. https://cran.r-project.org/web/packages/merTools/index.html (xaccessed June 26, 2019).

20 Zin CS, Chen LC, Knaggs RD. Changes in trends and pattern of strong opioid prescribing in primary care. Eur J Pain 2014; 18: 1343–51.

21 Department of Health. Controlled Drugs (Supervision of Management and Use) Regulations 2013: Information about the Regulations Title The Controlled Drugs (Supervision of Management and Use) Regulations 2013 Information about the Regulations. 2013 http://www.dh.gov.uk/publications (accessed July 17, 2019).

22 Fleming MF, Balousek SL, Klessig CL, Mundt MP, Brown DD. Substance Use Disorders in a Primary Care Sample Receiving Daily Opioid Therapy. J Pain 2007; 8: 573–82.

23 Park TW, Saitz R, Ganoczy D, Ilgen MA, Bohnert ASB. Benzodiazepine prescribing patterns and deaths from drug overdose among US veterans receiving opioid analgesics: case-cohort study. BMJ 2015; 350: h2698.

24 Gomes T, Juurlink DN, Antoniou T, Mamdani MM, Paterson JM, van den Brink W. Gabapentin, opioids, and the risk of opioid-related death: A population-based nested case–control study. PLOS Med 2017; 14: e1002396.

25 Brummett CM, Waljee JF, Goesling J, et al. New persistent opioid use after minor and major surgical procedures in us adults. JAMA Surg 2017; 152. DOI:10.1001/jamasurg.2017.0504.

26 Brat GA, Agniel D, Beam A, et al. Postsurgical prescriptions for opioid naive patients and association with overdose and misuse: retrospective cohort study. BMJ 2018; : j5790.

27 Faculty of Pain Medicine. Opioids Aware: A resource for patients and healthcare professionals to support prescribing of opioid medicines for pain | The Royal College of Anaesthetists. 2018. http://www.fpm.ac.uk/faculty-of-pain-medicine/opioids-aware (accessed April 24, 2018).

28 Frank JW, Levy C, Matlock DD, et al. Patients’ perspectives on tapering of chronic opioid therapy: A qualitative study. Pain Med 2016; 17: 1838–47.

29 Barnett ML, Olenski AR, Jena AB. Opioid-Prescribing Patterns of Emergency Physicians and Risk of Long-Term Use. N Engl J Med 2017; 376: 663–73.

30 Bridges SL. Health Survery for England-2011, health, social care and lifestyles. Chapter 9: Chronic Pain. 2012. https://digital.nhs.uk/data-and-information/publications/statistical/health-survey-for-england/health-survey-for-england-2011-health-social-care-and-lifestyles.

